# The relationship between Segmental Assessment of Trunk Control and Gross Motor Function Classification System in children with cerebral palsy

**DOI:** 10.64898/2026.01.20.26344472

**Authors:** Tania E. Sakanaka, Penelope B. Butler, Ian D. Loram

## Abstract

**Aim:** To determine the mechanistic relationship between segmental trunk control in the neutral vertical posture (NVP), assessed using the Segmental Assessment of Trunk Control (SATCo), and the Gross Motor Function Classification System (GMFCS); and hence to identify the means to enhance function in children with cerebral palsy (CP).

**Method:** This cross-sectional study included 101 children with CP (34 female, 10y(3y8m), 1.32(0.27)m, 33.4(18.4)kg) classified across GMFCS Levels I-V and tested with SATCo. Association and variation between GMFCS Levels and SATCo results were examined.

**Results:** SATCo results differed significantly (p<.05) between GMFCS Levels in static, active and reactive tests of trunk control. As neuro-ability increases through GMFCS Levels V-I, ability to control the head and trunk in NVP increases (*ρ*(99)=-0.61 to -1,p<.0001) and variation in head and trunk control increases (*ρ*(3)=-0.9 to -1,p<.05).

**Interpretation:** SATCo provides mechanistic insights supporting its use following GMFCS. In severe CP, NVP control is minimal across all children. In mild CP, large variation in results shows that SATCo discriminates between the use of full trunk control from compensatory strategies to achieve function. For each GMFCS Level, SATCo identifies the training required to improve trunk control in NVP, thus improving functional performance and reducing long-term risk of deformity.

**What this paper adds:**

- SATCo results are related to GMFCS Levels, and complements GMFCS
- SATCo provides the mechanistic explanation for what is observed in GMFCS
- SATCo-GMFCS reveals if function is attained with trunk control or compensatory strategies
- Compensatory strategies often used in mild CP are not captured by GMFCS
- SATCo identifies the training required to improve function and reduce deformity risk

**Graphical abstract:** 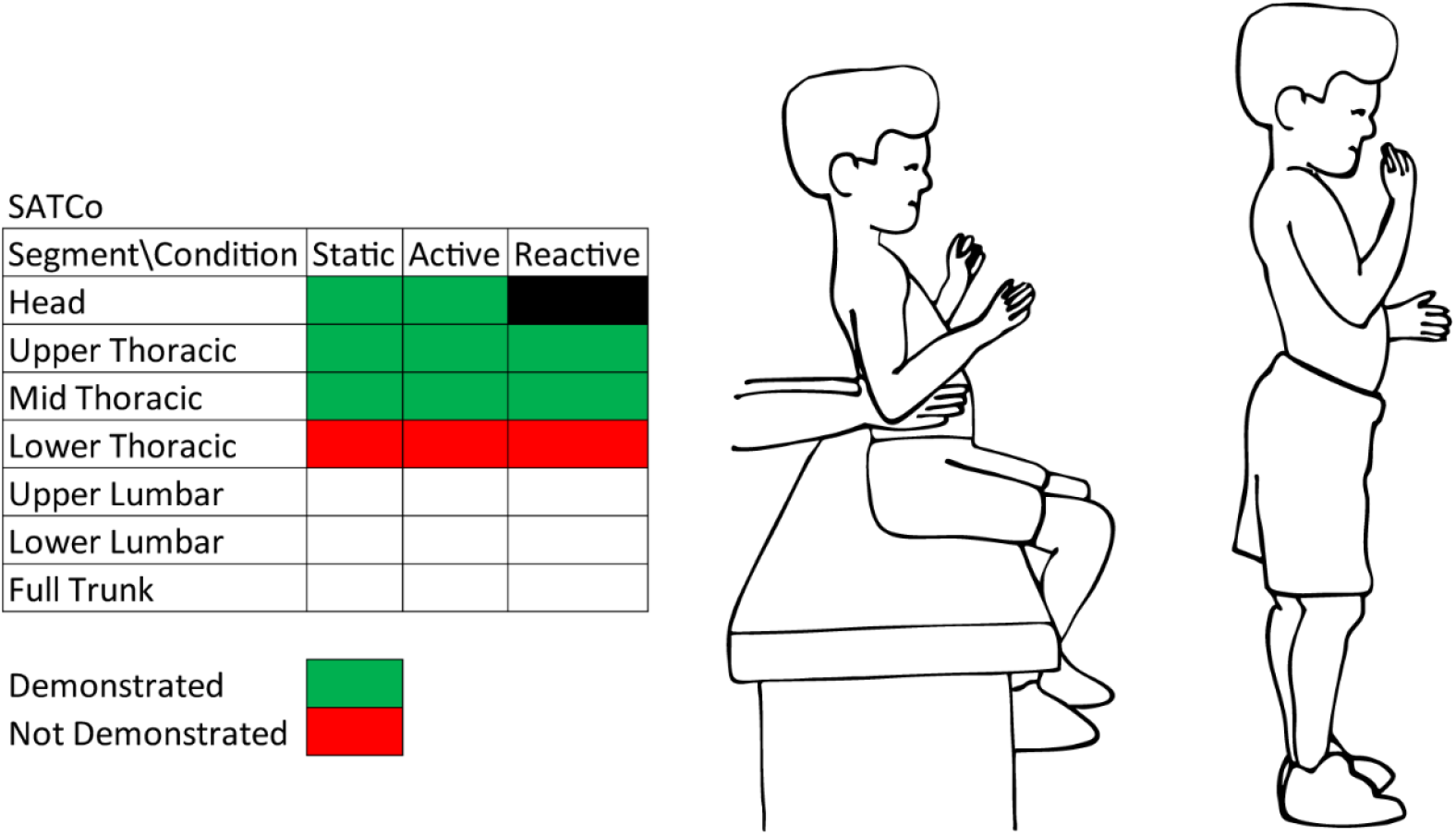

- Example above: GMFCS Level I child leaning backwards when tested for lower thoracic NVP trunk control. Same child showing compensatory lordotic lumbar posture while standing.
- SATCo can be used in combination with GMFCS to identify specific training targets to improve postural control, enhance function, and reduce deformity risk.

## Introduction

Cerebral palsy (CP) is a group of permanent movement and posture disorders caused by non-degenerative dysplasia or injury to the foetal or infant brain with no specific aetiology or a clear pathognomonic sign.^1^ Its classification in subtypes has been determined phenomenologically by various modalities of clinical evaluation or by functional classification systems (principally the Gross Motor Function Classification System – GMFCS^2,3^).^4^

GMFCS is an ordinal scale categorizing the gross motor function of children and adolescents with CP into five functional performance levels (I-V) across five age bands (<2 years, 2–4 years, 4–6 years, 6– 12 years, and 12–18 years), according to their ability to produce self-initiated movement and dependence on assistive devices^2^ (Table_1). Motor milestones are structured to reflect the developmental progression observed in typically developing (TD) children – from head control through running and jumping. Motor delays in children with CP mean that some milestones might never be reached.^5^ Children with greatest severity of disability (GMFCS V) may not achieve head control while a child classified at GMFCS II will achieve walking but run or jump with difficulty. GMFCS has strong stability^6–9^ and predictive validity, offering a valuable prognostic guideline for caregivers and clinicians.^7^

**Table 1.**
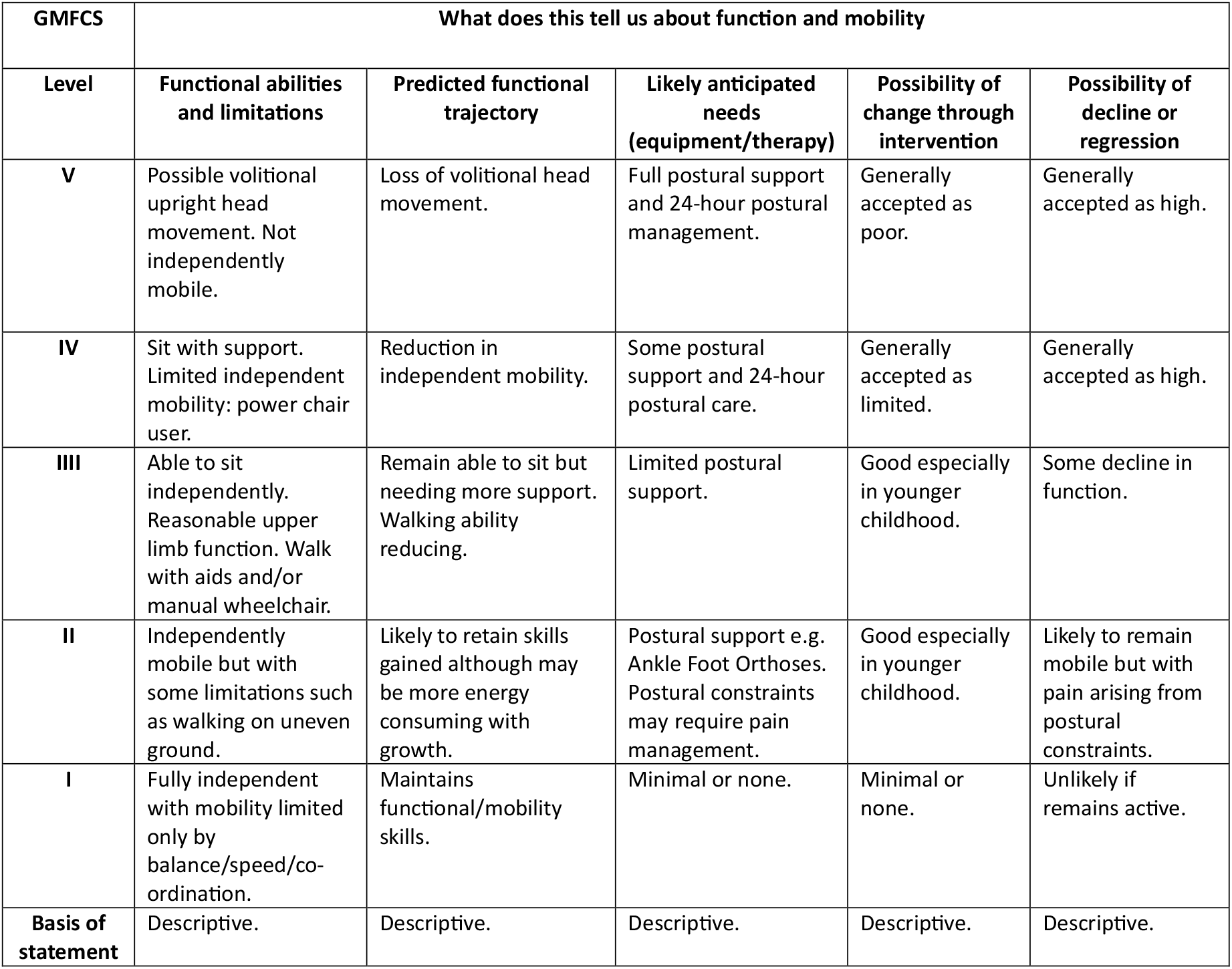
Description of GMFCS Levels.

The Segmental Assessment of Trunk Control (SATCo)^10^ assesses the developmental progression of head and trunk control in the neutral vertical posture (NVP).^11^ Head/trunk control in NVP provides the postural foundation for both upper and lower limb function and ambulation12^–14^. In TD children, achievement of independent sitting is linked to the cephalocaudal development of full trunk control in NVP.^15–17^ However, children with CP show delay or cessation of this cephalocaudal progression.^18^ This frequently leads to the use of compensatory strategies to achieve sufficient stability to function, for example bracing the hands on the seat to enable stability. Once established, these compensations can limit further functional development since the basis of NVP is lacking.

Although based on mechanistic principles of postural control, SATCo is clinically descriptive. SATCo considers six defined head and trunk segments, as well as full trunk control, under static, active, and reactive conditions: SATCo identifies **the trunk segment at which the child is acquiring NVP control**. i.e. the highest trunk segment at which independent NVP control is not maintained (Table_2).

**Table 2.**
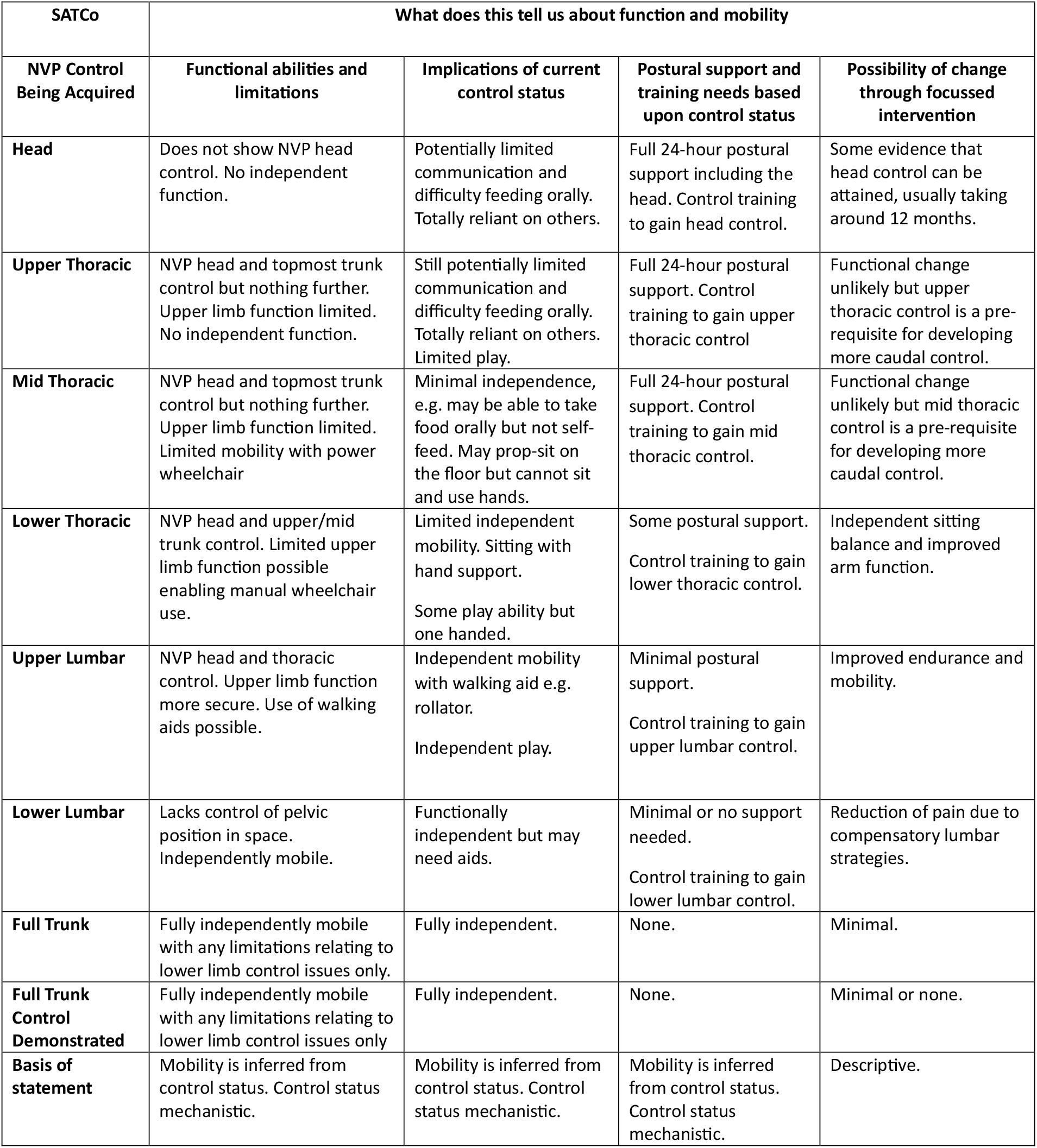
Description of SATCo Segments.

The GMFCS classifies usual motor performance, describing what children with CP are able to do without addressing the quality or mechanisms of movement. This study explored the relationship between the mechanistic information provided by SATCo and the descriptive information offered by the GMFCS and how these two tests might complement each other.

## Method

### Study design, setting, and participants

This study reports the subset of children with CP from a large observational cross-sectional study aimed at improving objectivity of the assessment by developing a clinical tool with live imaging analysis technology to automate SATCo analysis (UK Medical Research Council-funded project “Quantification of head and trunk control for children with neuromotor and neuromuscular disorders”, MR/T002034/1). This dataset was subject to thorough quality control performed by the creator of SATCo (PBB, author of this study) and a rigorous standard of test procedure and scoring, since this dataset is being prepared to train an objective SATCo (oSATCo) AI system.

Study information sheets were provided to parents/guardians with opportunity to ask questions before signing a written informed consent. The London-Brent Research Ethics Committee (IRAS project ID: 233469) granted ethical approval.

Inclusion criteria were children with a clinical diagnosis of CP between 2 and 16 years old where both parents/guardians had sufficient understanding of English to give informed consent. Children who posed a physical risk during testing to themselves, or the research team were excluded. Between 2022-2024, local community paediatric physiotherapists screened for eligibility children from their services resulting in recruitment of a convenience sample of 101 children (Table 1). Children with fixed spinal deformities were included but no SATCo outcomes below commencement of the deformity were reported.

Data collected from participants included body height and weight, sex, and spine status relating to deformities. GMFCS Level classification had been undertaken previously by the local teams.

### Experimental protocol

Three Intel® RealSense™ Depth Cameras D435 were used to record SATCo. The child was seated on a Leckey Therapy Bench (James Leckey Design Ltd, Lisburn, N. Ireland), his/her hip and knee flexed at 90° and pelvis secured in NVP. Two trained assessors conducted SATCo with sessions lasting 10-40 min depending on performance (Figure 1).

**Figure 1.**
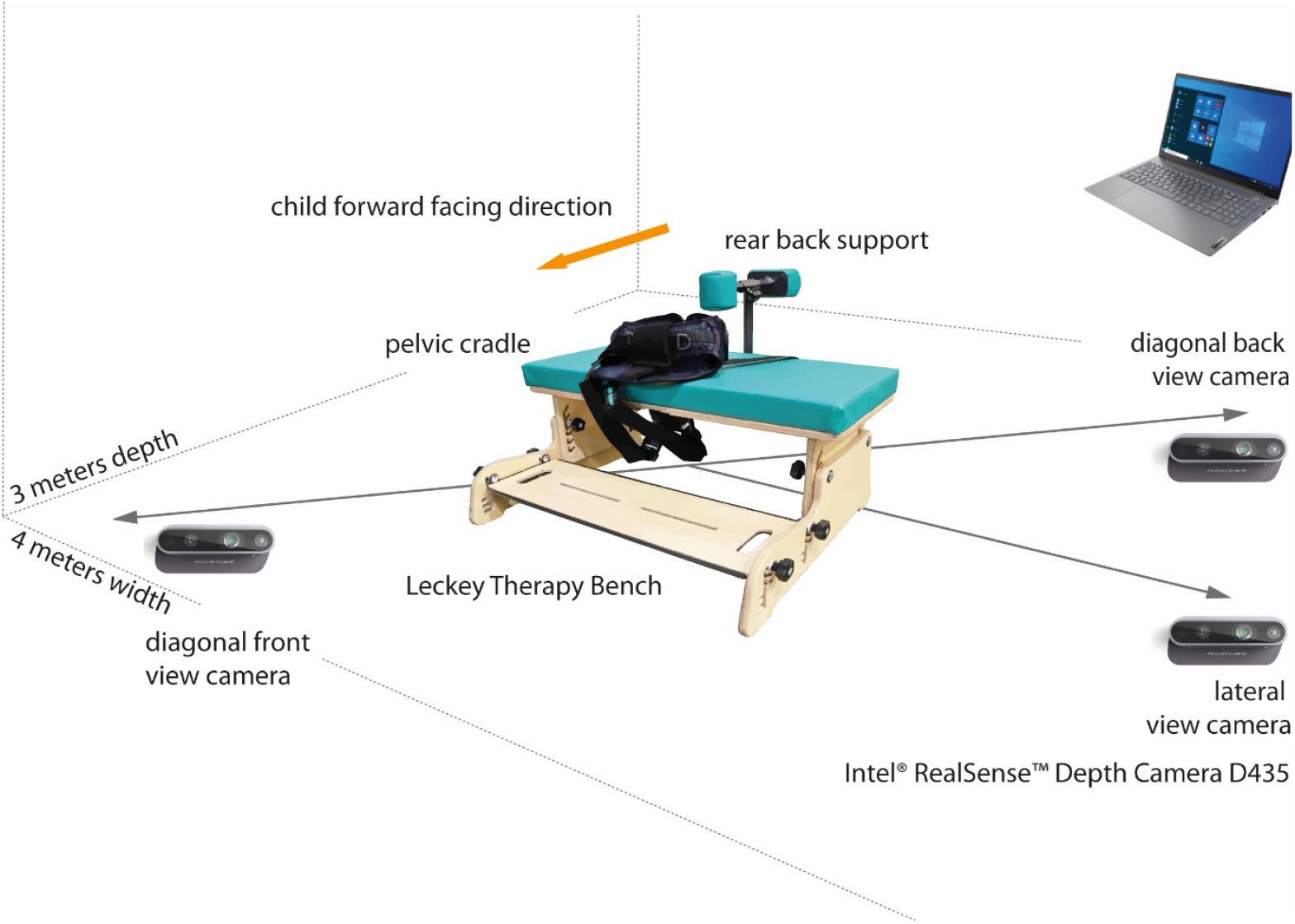
Experimental setup. The room used for recording was minimum 3 by 4 meters. Cameras were adjusted to record lateral, diagonal front and back views of the child’s posture. Camera height and distance was adjusted according to each child’s size.

The test was conducted and visually NVP-checked by the first assessor. Manual horizontal support for each trunk segment was given by the second assessor, for *head* – support at the shoulder girdle, *upper thoracic* – axillae, *mid thoracic* – inferior scapula, *lower thoracic* – over lower ribs, *upper lumbar* – below ribs, *lower lumbar* – pelvis, and *full trunk* – no support given. *Static* (unsupported NVP for 5 seconds), *active* (NVP during a child-initiated head turn to left and right), *reactive* (ability to return to NVP following a nudge from the front, back and both sides, not tested at the *head* segment) forms of control were tested at each segment.

Control was defined as *demonstrated, not demonstrated*, or *not tested* for each segment and each form of test. Demonstration of control was confirmed if the child maintained the unsupported trunk segments above the manual support in NVP. Control was considered *not tested* if there was an assessor error.

SATCo tests were scored by PBB and TES, authors, and TP, contributor to this study. Quality control was performed by PBB, creator of SATCo with more than 30 years of experience working with children with cerebral palsy.

### Statistical analysis

IBM® SPSS® Statistics for Windows (v.28, IBM, USA) and RStudio: Integrated Development for R (v.2024.04.2+764, Posit Software, USA) were used for statistical analysis. Mean and standard deviation from body height, weight, and age from children in each GMFCS Level were used to characterize the sample. SATCo outcomes (the highest segment at which control was being attained) for each form of control (static, active, reactive) were transformed in ranks (*control not demonstrated: head*=1, *upper_thoracic*=2, *mid_thoracic*=3, *lower_thoracic*=4, *upper_lumbar*=5, *lower_lumbar*=6, *full_trunk*=7; *control demonstrated: full_trunk_demonstrated*=8), then checked for normality with Shapiro-Wilk test. Initially, differences in SATCo results medians between GMFCS Levels were calculated with Kruskal-Wallis H test. Post-hoc pairwise comparisons were performed with Dunn’s test. Levene’s test for equality of variances was used to check if variances in SATCo results differed between GMFCS Levels with sample variances calculated for each subgroup. Finally, Spearman’s rank correlation coefficient (Spearman’s ρ) test was used to verify any association between SATCo outcome medians and variances versus GMFCS Levels. Spearman’s test results were ranked as weak between ±0.1 and ±0.3, moderate between ±0.4 and ±0.6, strong between ±0.7 and ±0.9, and perfect when ±1.0^19^. P-values were considered significant at 0.05.

## Results

Table_3 details the 101 children with CP recruited for this study.

**Table 3.** Clinical and anthropometric characteristics of participants grouped by GMFCS Level.

| GMFCS Level | Sex | Age (Years, Months)<br>mean (SD) | Body height<br>(cm) | Body weight<br>(kg) | SCPE classification |
| --- | --- | --- | --- | --- | --- |
| V<br>(n=13) | 5 F<br>8 M | 8 years, 10 months<br>(2 years, 10 months) | 118.8 (15.8) | 20.8 (6) | 7 Spastic bilateral<br>6 Dyskinetic |
| IV<br>(n=29) | 12 F<br>17 M | 9 years, 7 months<br>(3 years, 6 months) | 126.9 (20.8) | 30.8 (19.5) | 1 Spastic unilateral<br>21 Spastic bilateral<br>7 Dyskinetic |
| III<br>(n=12) | 6 F<br>6 M | 10 years, 2 months<br>(4 years, 3 months) | 125.1 (19.4) | 34.6 (16.7) | 2 Spastic unilateral<br>9 Spastic bilateral<br>1 Dyskinetic |
| II<br>(n=21) | 5 F<br>16 M | 9 years, 6 months<br>(3 years, 10 months) | 133 (21.7) | 32.3 (14.7) | 10 Spastic unilateral<br>10 Spastic bilateral<br>1 Ataxic |
| I<br>(n=26) | 6 F<br>20 M | 11 years, 2 months<br>(4 years, 1 month) | 146.2 (22.4) | 43.3 (21.5) | 21 Spastic unilateral<br>5 Spastic bilateral |
| Average of all<br>participants<br>(n=101) | 34 Female<br>67 Male | 10 years, 0 months<br>(3 years, 8 months) | 132.2(22.6) | 33.4(18.4) | 34 Spastic unilateral<br>52 Spastic bilateral<br>14 Dyskinetic<br>1 Ataxic |

Visual representation of the results is shown in Figure_2 (static), Figure_3 (active), and Figure_4 (reactive control). Static SATCo results moved progressively caudally from GMFCS Level V to Level I, ranging from children still acquiring NVP head control to children with full trunk control who could run and jump. Children with greater disability (GMFCS III-V) did not show active or reactive NVP control beyond head (active) or upper thoracic (reactive) segments while children of GMFCS Levels I and II showed active and reactive control comparable to their static control.

**Figure 2:**
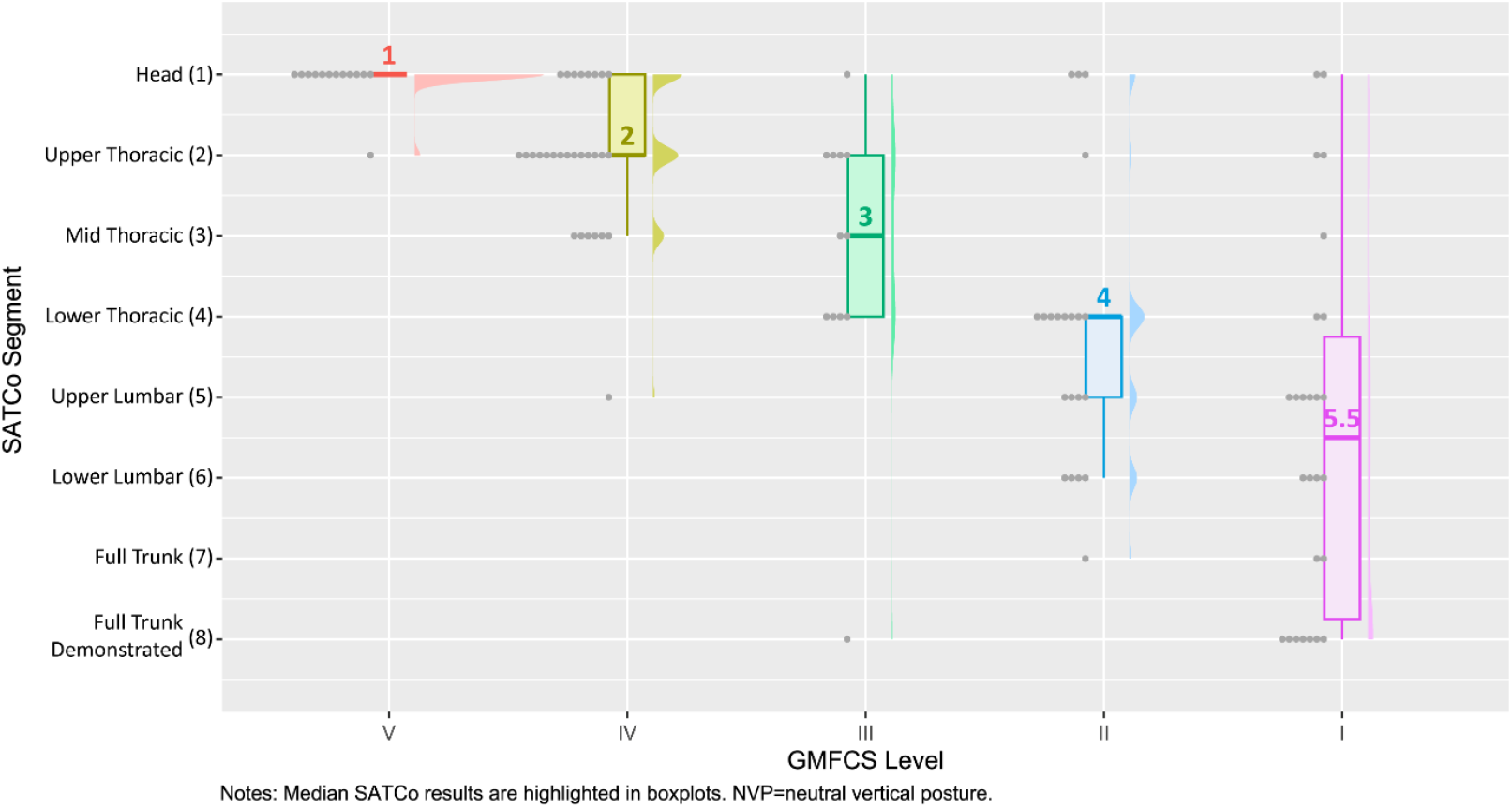
Head and trunk NVP static control in children with CP. Raincloud plot showing distribution of SATCo results in static control for different GMFCS Levels in children with cerebral palsy

**Figure 3:**
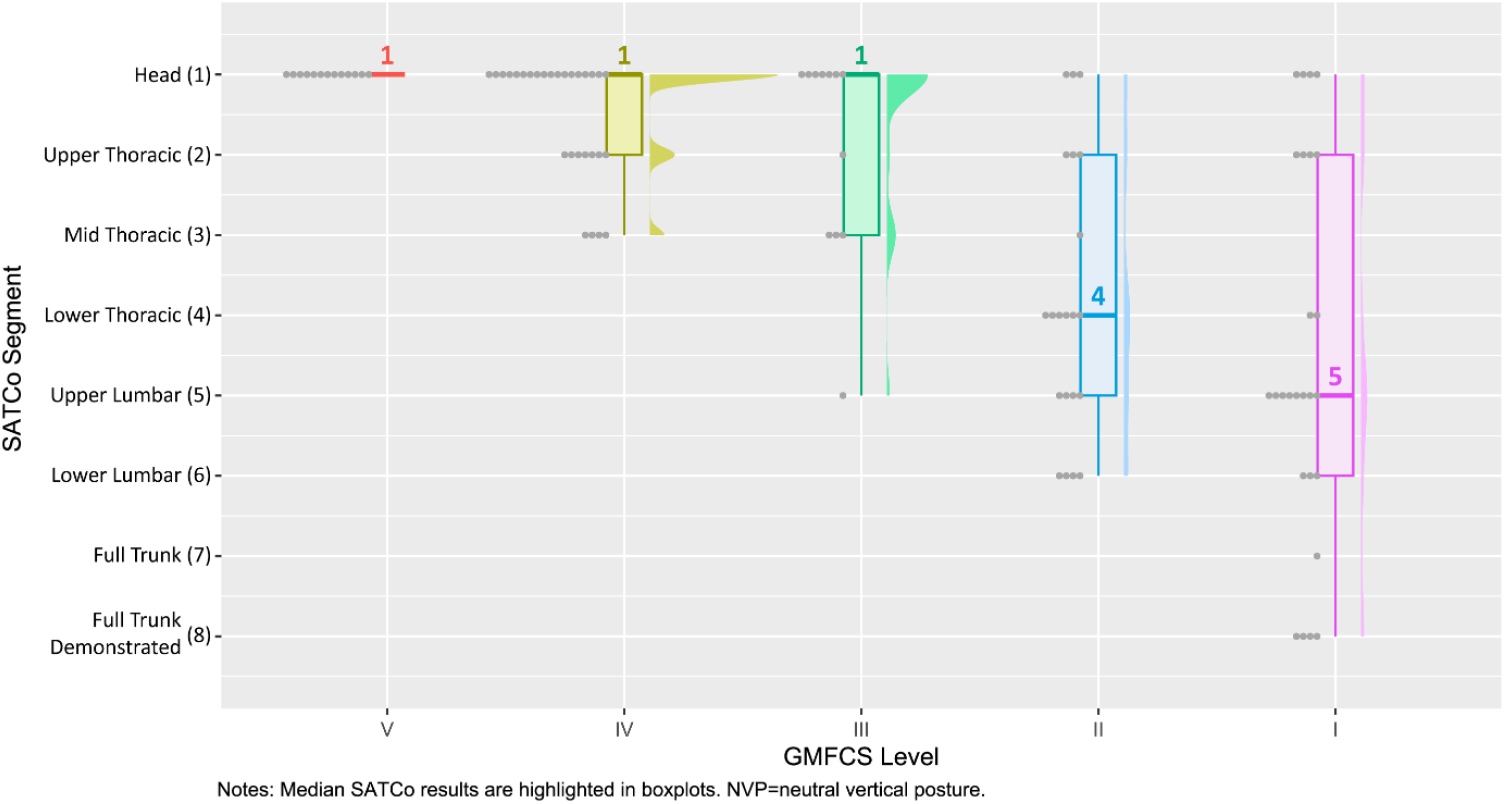
Head and trunk NVP active control in children with CP. Raincloud plot showing distribution of SATCo results in active control for different GMFCS Levels in children with cerebral palsy

**Figure 4:**
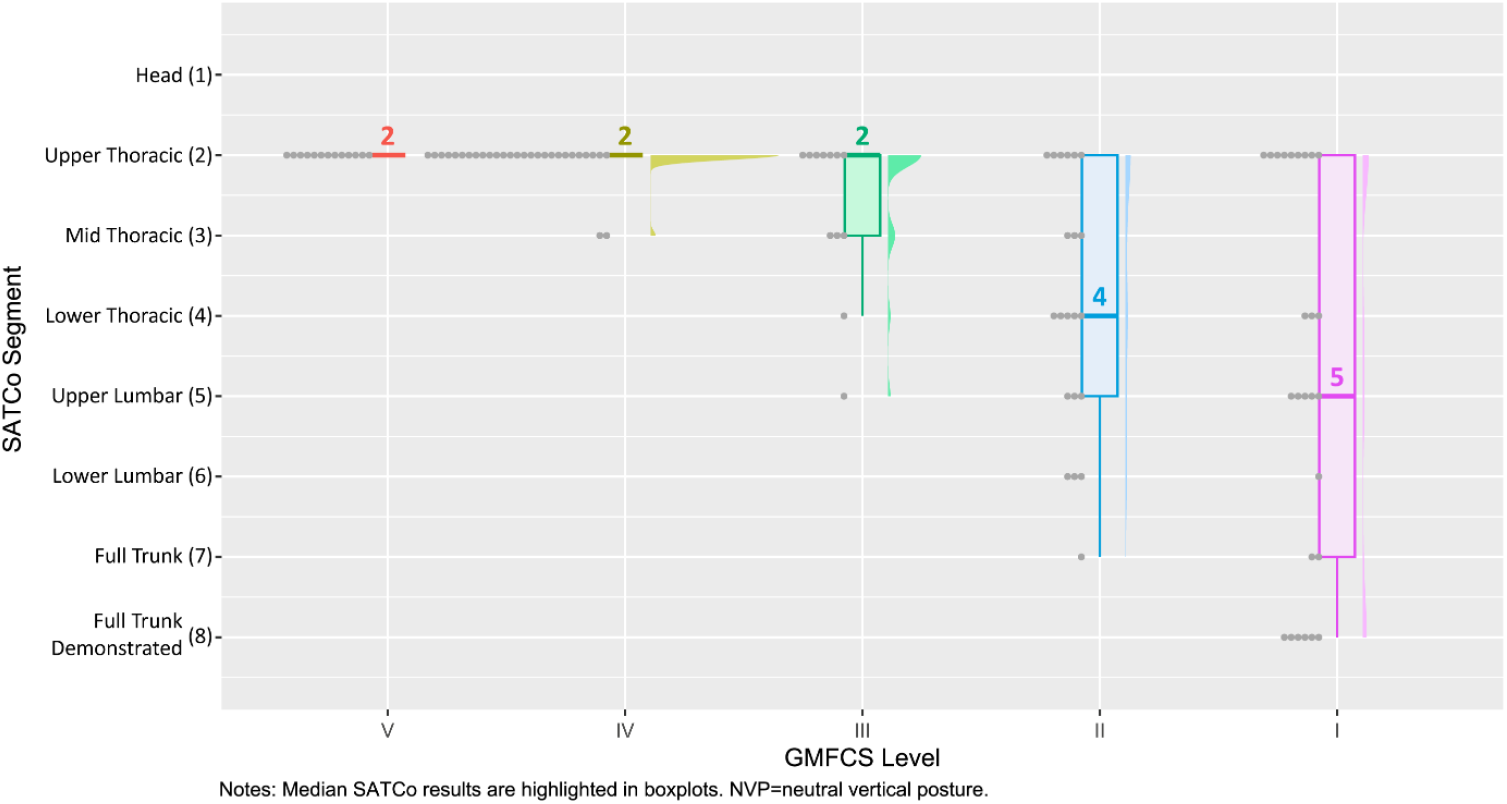
Head and trunk NVP reactive control in children with CP. Raincloud plot showing distribution of SATCo results in reactive control for different GMFCS Levels in children with cerebral pals

All children at GMFCS Level V (except one) were acquiring static head control while children at GMFCS Level IV had gained static head control and were acquiring upper thoracic static control. Children at GMFCS Level IV did not demonstrate active head or upper thoracic control or reactive upper thoracic control.

Children at GMFCS Level III were acquiring static mid thoracic control but greater variability was shown ranging from acquiring head to full trunk control demonstrated. However, results for active and reactive control remained at head and upper thoracic segments.

At GMFCS Levels II and I even greater variability with more evenly distributed results was observed. Scores for active and reactive control more closely matched those for static control, in contrast to children at Levels V through III.

Findings for children at GMFCS Level II indicated that they were acquiring static, active and reactive lower thoracic control while children at GMFCS Level I were, on average, acquiring upper lumbar control.

Median and variance results for each subgroup are shown in Table_4.

**Table 4.** SATCo outcomes in different GMFCS Levels and types of control.

| GMFCS Level | Static |  |  | Active |  |  | Reactive |  |  |
| --- | --- | --- | --- | --- | --- | --- | --- | --- | --- |
|  | Median | s<sup>2</sup> | Range | Median | s<sup>2</sup> | Range | Median | s<sup>2</sup> | Range |
| V (n=13) | 1 | 0.08 | 1-2 | 1 | 0 | 1 | 2 | 0 | 2 |
| IV (n=29) | 2 | 0.82 | 1-5 | 1 | 0.54 | 1-3 | 2 | 0.07 | 2-3 |
| III (n=12) | 3 | 3.30 | 1-8 | 1 | 1.72 | 1-5 | 2 | 0.97 | 2-5 |
| II (n=21) | 4 | 2.96 | 1-7 | 4 | 2.96 | 1-6 | 4 | 2.53 | 2-7 |
| I (n=26) | 5.5 | 5.05 | 1-8 | 5 | 5.54 | 1-8 | 5 | 5.80 | 2-8 |

Notes: Median and variance are not adjusted for different sample sizes in each GMFCS Level. s<sup>2</sup> = sample variance.

Median SATCo results varied between GMFCS Levels, in static, active and reactive control (Table_5A, p<0.0001). Spearman’s test revealed negative association between median SATCo results and GMFCS Levels (Table_5D, p<0.0001), ranked as good for static, and moderate for active and reactive control. As neuro-disability decreased from GMFCS Levels V through I, progressively more head/trunk segments were controlled in NVP across static, active, and reactive control.

**Table 5.**
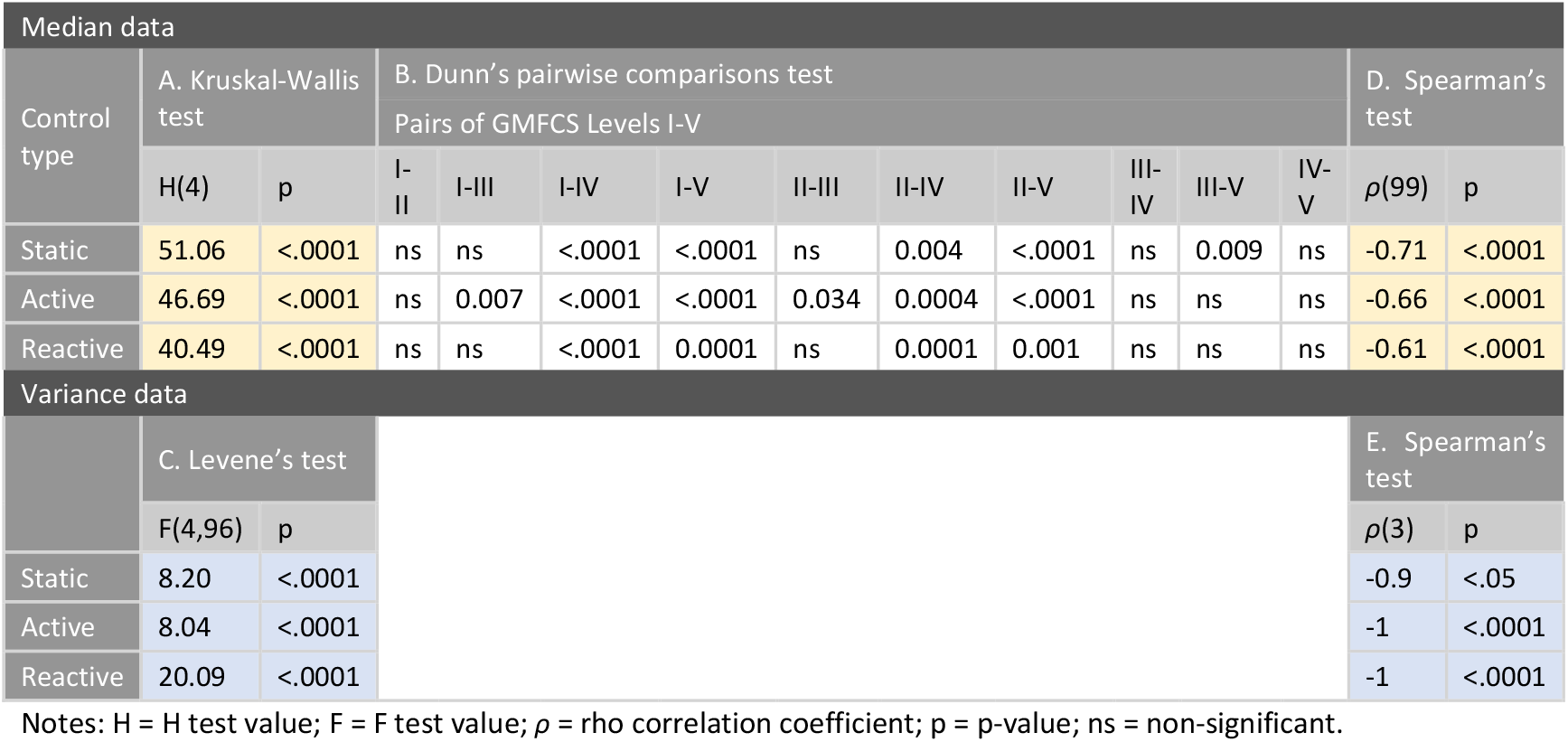
Results from statistical analysis. (A) and (B) Differences in SATCo medians between GMFCS Levels: Kruskal-Wallis test and post hoc Dunn’s pairwise comparisons test revealed a difference in median SATCo results between GMFCS Levels, in static, active and reactive control. (C) Differences in SATCo variances between GMFCS Levels: Levene’s test showing no homogeneity in variances of SATCo results between GFMCS Levels. (D) and (E) Association between SATCo medians and variances versus GMFCS Levels: Spearman’s rank correlation coefficient test revealed negative association between median and variance SATCo results and GMFCS Levels.

Variances of SATCo results varied between GFMCS Levels, in static, active and reactive control (Table_5C, p<0.0001). Spearman’s test revealed a negative association between variances in SATCo results and GMFCS Levels (Table_5E, p<0.05), good in static, and perfect in active and reactive control. As neuro-disability decreased from GMFCS Levels V through I, variability in the number of head/trunk segments controlled in NVP increased across static, active, and reactive control.

## Discussion

### SATCo outcomes are related to GMFCS Levels

In 101 children with CP, this cross-sectional study compared segmental assessment of head/trunk control (SATCo), with usual motor performance classified by GMFCS. SATCo outcomes showed a clear, negative correlation with GMFCS Levels: NVP control-demonstrated progressed to more caudal segments as neuro-disability decreased. Also, variability in NVP control-demonstrated increased as neuro-disability decreased. Children at Level V showed minimal variance and almost no NVP control whereas children at GMFCS Level I ranged from still acquiring NVP head control to demonstrating full NVP trunk control.

### SATCo provides the mechanistic explanation for what is observed in GMFCS and how function may be improved

The majority of children at GMFCS Level V lacked NVP head control (static and active) (Figures_2-4). These children may be learning head control but, as yet, remain unable to maintain NVP (Table_2). Consistent with Level V GMFCS classification, these children are totally dependent and require 24-hour postural support (Table_1). GMFCS classification acknowledges volitional upright head movement but not necessarily in NVP. The ability to control the head in NVP has a profound impact on these children’s capacity to interact with others, engage with their environment and begin to communicate such as by eye-pointing. Research indicates that following a SATCo outcome of head control not demonstrated, head control can be achieved using therapy focused on improving NVP control (Targeted Training, TT)^20,21^.

Children at GMFCS Level IV were generally acquiring upper thoracic static NVP control i.e. they had acquired head control previously (Table_2). These children will still require 24-hour postural support but may be able to use a power wheelchair for mobility (Table_1). Active control involves the ability to move the head while maintaining NVP, while reactive control requires the ability to return the head to NVP following perturbation: results demonstrated that both skills had yet to be acquired. This thoracic control status impacts upper limb function, consistent with the GMFCS Level IV description. To achieve functional abilities comparable to those seen at GMFCS Level III, these children would need to develop upper and mid thoracic control and be progressing toward lower thoracic control.

The 12 children at GMFCS Level III were, on average, learning static mid thoracic control. These children are generally more mobile and able to sit with hand support, walk with aids or use a manual wheelchair. This need for hand support is clarified by SATCo findings of children still acquiring mid thoracic control. SATCo can be used alongside GMFCS to guide interventions to improve trunk control that could potentially reduce reliance on hand support for mobility.

Children at GMFCS Level II were acquiring lower thoracic static, active, and reactive control. They are able to walk independently with minimal limitations (Table_1). However, there is much greater variability in SATCo result, with some children lacking full head control while others were acquiring full trunk control. Although upper thoracic reactive control was not demonstrated in some children, they had sufficient more caudal static and active control to function. These more caudal control elements enable their use of compensatory strategies to achieve function. This contrasts with children of GMFCS Levels V and VI where their trunk control limitations preclude the use of compensatory strategies, thus impacting function. Of the 21 children at GMFCS Level II, 12, 13, and 14 (for static, active, and reactive control, respectively) did not demonstrate lumbar control. A common compensation in this group is to lock the lumbar spine in full extension (lumbar lordosis), negating the need for active lumbar spine control. This compensation enables independent walking but may contribute to their difficulty on uneven terrain. However, this compensatory pattern may also contribute to long-term problems including lumbar pain and degenerative changes.

Out of 26 children classified at GMFCS Level I, 19, 16, and 14 (for static, active, and reactive control, respectively) were still acquiring upper or lower lumbar control. It is notable that, although classified as ‘fully functional’ with walking limited only by balance, speed, or coordination (Table_1), full trunk control was not necessarily present (Figure_5). Where trunk control deficits exist, compensatory strategies are likely to be used to achieve function. Typically, these children adopted the hyper-lordotic lumbar posture seen in children of GMFCS Level II with consequent pain and development of spinal deformities often reported in severe cases^22^ but not usually reported in mild cases.

**Figure 5.**
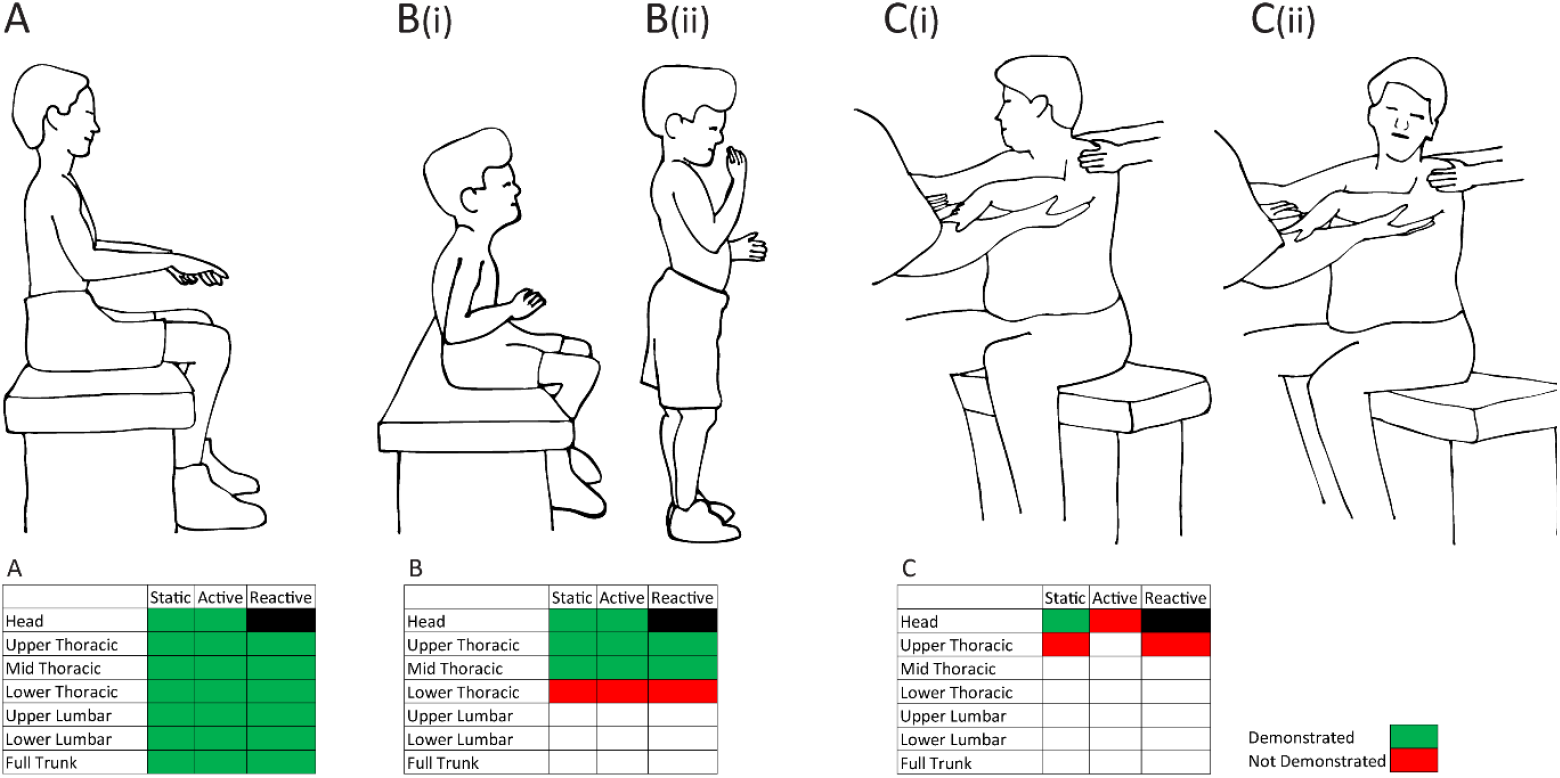
Variation in results in children at GMFCS Level I. A. Child showing full NVP trunk control. B(i). Child using slumped sitting posture and unable to sit in NVP. B(ii) Same child showing compensatory lordotic lumbar posture while standing. C(i). Child being tested for active head control turning head to right. C(ii). Same child showing no NVP head control while turning the head to the left during active head control. SATCo results are shown below images.

Two children classified at GMFCS Level I did not demonstrate full head control. They were unable to maintain the head in NVP but, nevertheless, had sufficient trunk control to walk. While postural anomaly may be inconsequential in childhood and adolescence, it could lead to future degenerative changes and cervical spine pain in the future. Trunk control training may help reduce long-term problems.

In summary, SATCo identifies the underlying control status that results in the functional abilities demonstrated. This clarifies a potential ‘route forward’ through focussed control training leading to greater functional abilities.

Other authors have explored the relationship between trunk control and function. Heyrman et al. (2013)^23^ used the Trunk Control Measurement Scale (TCMS) and motion capture analysis and found that greater trunk movement during gait was related to more trunk deficits. The majority of their cohort of 20 children with CP at GMFCS Levels I and II demonstrated full static trunk control (median 20/20). The contrast with the findings of the present study relate to the differing underlying concept of SATCo and TCMS and the differing specificity of the tests. TCMS assesses overall trunk posture during functional movements including reaching but does not focus on the NVP. It also requires a child to use hand support, thus not testing trunk control *per se*. While the SATCo results of the present study supports Heyrman et al. (2013)’s finding that trunk control deficits are present in children at GMFCS Levels I and II who can walk, SATCo additionally reveals the precise location of the trunk control deficits.

### Is there room for change? Clinical implications of identifying compensatory strategies

The functional trajectory through life is determined both by the original insult to the brain and by the input received by the child, such as physiotherapy/surgery. Some recent studies on GMFCS stability confirmed that the classification is less stable in children under 4/6 years old.^9,24,25^ The number of physiotherapy sessions received by children with spastic quadriplegia in the 2-4 year age range and with a lower GMFCS classification (higher ability) impacted the amount of change.^25^

Since the biomechanical input is very different between reclined/supported postures such as lying and rolling and upright postures such as sitting/standing, it is very unlikely that (physio)therapy strategies in non-vertical postures will result in improvement in upright postural skills/function. In the upright posture, control is gained from head downwards and thus any physiotherapy intervention that uses this process is more likely to be effective and has been demonstrated in children with CP.^26–29^

As an example, the therapy strategy linked with SATCo is Targeted Training (TT). In TT the child is supported directly below the identified targeted segment i.e. the highest head/trunk segment at which NVP control is not demonstrated, and through play and social interaction, the child is engaged to use the free portion of their head/trunk and upper limbs (not for head control training). As NVP control is gained, the support is moved segmentally caudally. This simplifies control learning to one head/trunk segment, with those segments above already demonstrating or having gained NVP control.^28^ The increase in NVP control provides the ‘building blocks’ for function to emerge and is a therapy strategy that can be clinic or home based.^30^ Such therapies provide a basis for future trunk control therapy strategies evolving from the results provided by this study.

### Strengths and limitations

The reported data is a subset of a larger study (MR/T002034/1) creating a clinically based neural network tool that will provide automated objective analysis and feedback for SATCo testing. The tool will enable clarity and facilitate SATCo training/delivery. A rigorous standard for test conduct and scoring was set to ensure that all data encoded to this tool was consistent to train the neural network appropriately. A quality control process led to rigorous analysis resolving edge cases.

This study benefits from a large, heterogeneous sample spanning all GMFCS Levels and rigorous application of SATCo to isolate segmental postural capacity. However, its cross-sectional design limits causal inference, and variability in tester experience may have influenced SATCo outcomes.

### Conclusion and future studies

SATCo offers mechanistic insights that explain the functional limitations observed in children with CP. Following a GMFCS classification, SATCo is a logical next step taking 15 minutes to 30 minutes with the child. SATCo complements the GMFCS by providing information about segmental trunk control in NVP. GMFCS remains essential for describing functional classification and predicting broad service needs, whereas SATCo provides segment-specific insight into postural status and possible potential for change. Using both measures clarifies where functional performance is affected by trunk control status and where to target intervention.

## Data Availability

All data produced in the present work are contained in the manuscript.

## Abbreviations

CP: Cerebral palsy
GMFCS: Gross Motor Function Classification System
NV, NVP: Neutral vertical, neutral vertical posture
SATCo, oSATCo: Segmental Assessment of Trunk Control, Objective Segmental Assessment of Trunk Control
TCMS: Trunk Control Measurement Scale
TD: Typically developing
TT: Targeted Training

## Acknowledgments

We gratefully acknowledge Teresa Perez and Natalia Rajkowska from Manchester Metropolitan University. We also thank James Leckey Design Ltd (Lisburn, Ireland) for kindly providing Leckey Therapy Benches for this study.

## Funding

This research was funded by the Medical Research Council (MRC) (MR/T002034/1).

## Notes

### Competing Interest Statement

The authors have declared no competing interest.

### Author Declarations

The London-Brent Research Ethics Committee (IRAS project ID: 233469) gave ethical approval for this work.

### Summary of Updates

(1) Caption for the graphical abstract, from -Example to the left- to -Example above- in page 3. (2) Added -(Figure_5)- to -Out of 26 children classified at GMFCS Level I, 19, 16, and 14 (for static, active, and reactive control, respectively) were still acquiring upper or lower lumbar control. It is notable that, although classified as fully functional with walking limited only by balance, speed, or coordination (Table_1), full trunk control was not necessarily present (Figure_5).- in page 12. (3) Added page numbers.

